# Dynamical survival analysis for epidemic modeling under partial observability: A case study of COVID-19 in Kenya

**DOI:** 10.1101/2025.09.30.25337023

**Authors:** Josephine Wairimu, Andrew Gothard, Grzegorz Rempala, Bosueng Choi, the Lorem Ipsum Consortium

## Abstract

We apply the Dynamical Survival Analysis (DSA) framework to derive an individual-level epidemic model suitable for analyzing partially observed data in both longitudinal and cross-sectional formats. Within this framework, survival and hazard functions are constructed from available random samples of infection and recovery times, enabling principled, likelihood-based inference. To demonstrate the utility of this approach, we analyze the COVID-19 outbreak in Kenya from 2020 to 2022, focusing on three of the five observed pandemic waves—the first, second, and the most severe, fifth wave. The model is estimated using the Hamiltonian Monte Carlo method implemented in Stan. Results indicate a good fit to the observed epidemic curves and reveal substantial variation in response times for patient identification and isolation across different waves. These findings underscore the flexibility of DSA as a novel inference tool for epidemic modeling under partial observability.

## Introduction: The COVID-19 Outbreak in Kenya

The COVID-19 pandemic in Kenya was part of the global outbreak of coronavirus disease in 2019 (COVID-19), caused by the severe acute respiratory syndrome coronavirus 2 (SARS-CoV-2). The first confirmed case in Kenya was reported on March 12, 2020, in the capital city, Nairobi. The initial cases were also detected in the coastal area of Mombasa. As of late 2024, Kenya had reported over 344,000 confirmed cases and approximately 5,689 deaths. However, the true figures are widely believed to be considerably higher due to underreporting. The pandemic significantly strained the country’s healthcare system, affecting the delivery of essential health services. The Kenyan government implemented a range of measures to curb the spread of the virus, including lockdowns, curfews, travel restrictions, and the promotion of hygiene practices such as handwashing and mask-wearing. The country launched its COVID-19 vaccination campaign in early 2021. By early 2025, more than 14 million vaccine doses had been administered, with over 11 million people fully vaccinated. These efforts are believed to have contributed significantly to the subsequent decline in infections. The pandemic has had profound socioeconomic effects in Kenya, including job losses, reduced income, and higher poverty levels. The informal sector, which employs a significant portion of the population, was particularly hard hit. Education was also disrupted, with schools closing for extended periods and transitioning to remote learning, which presented challenges due to limited access to digital resources. This digital divide widened the gap between rich and poor students. Innovations in healthcare delivery, such as telemedicine and mobile health services, have been accelerated by the pandemic, improving access to care in remote areas [1].

Mathematical models have been used to describe the spread of COVID-19 in Kenya focusing on epidemiological trends, the effectiveness of interventions, and the socio-economic effects of the pandemic [2]. These models have been instrumental in guiding public health interventions and understanding the spread of COVID-19 in Kenya. For instance, Mbogo and co-authors developed a Susceptible–Exposed–Infectious –Hospitalized–Quarantined–Recovered–Dead (SEIHQRD) model to compute the basic reproduction number ℛ_0_ and to assess the key factors driving COVID-19 transmission in Kenya. Their findings indicated that long-term non-pharmaceutical interventions were essential for epidemic control and highlighted the importance of specialized medical care for individuals with comorbidities [3, 4]. Building on these insights, subsequent studies explored the spatial and demographic dimensions of the epidemic in Kenya, integrating mobility and population structure into transmission models. In particular, Samuel and co-authors simulated SARS-CoV-2 transmission within and between various Kenyan regions and age groups. Their model incorporated human mobility data, national census data, and Kenya-specific social interaction rates to demonstrate that asymptomatic individuals played a substantial role in shaping the epidemic trajectory [5]. In the same paper, Samuel and colleagues also developed a mathematical model to forecast the potential incidence and overall magnitude of the COVID-19 epidemic in Kenya. Their approach adapted key epidemiological parameters from the initial COVID-19 outbreak in China to the Kenyan context [5]. A model by [6] used a deterministic approach to understand the dynamics of COVID-19 infection among the Kenyan population with a vital interest in two proportions; people with other chronic illnesses and those who do not have other chronic illnesses. The study found out that that low adherence to the measures put in place to curb the disease increases infection in the population. Hospitalization and home-based care programs show that an increased rate of hospitalization and care lowers infection.

The purpose of this paper is to examine the spread of COVID-19 in Kenya through the lens of simplified mathematical models. The pandemic has underscored the importance of robust public health modeling to generate accurate predictions and help mitigate disruptions such as those experienced in Kenya. To improve model accuracy for both historical and future pandemics, we develop models that extend classical SIR dynamics by incorporating techniques from the Dynamical Survival Analysis (DSA) [7, 8] and the Poisson contact process [9].

The remainder of the paper is structured as follows: in Section 2, we introduce and derive the DSA framework, including the survival and hazard functions. In Section 2.2, we formulate the likelihood function. In Section 3, we presents our data analysis, where we fit the infection curve of the model to histogram data from three major COVID-19 waves in Kenya. Finally, we offer a discussion of the findings in Section 4

### 1 Dynamical Survival Analysis (DSA)

To study and predict the spread of infectious diseases or the dynamics of ecosystems, mathematical modelers often use compartmental models to classify individuals based on their epidemiological status. These models provide a powerful framework for understanding complex systems by simulating the movement of individuals or substances between distinct compartments within a population [10].

In the original Kermack-McKendrick SIR model, the population is divided into susceptible (S), infected (I) and recovered/removed (R) compartments. The evolutions of the population averages in compartments with time are defined as *s*_*t*_, *ι*_*t*_, and *r*_*t*_ and the system of ordinary differential equations (ODEs)

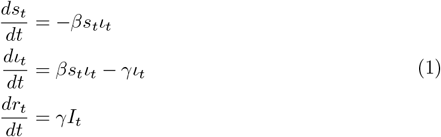

where (*β*) is the transmission rate and (*γ*) is the removal rate [11]. This simple compartmental model has been applied to a wide range of epidemic outbreaks to predict the epidemic trajectory, estimate key epidemiological parameters like the basic reproduction number (ℛ_0_ = *β/γ*), a threshold parameter which represents the average growth rate of secondary cases in a completely susceptible population [12, 13], and evaluate the impact of public health interventions.

The limitations of ordinary differential equation (ODE) models stem from their reliance on averaged individual dynamics, which makes it difficult to capture the stochastic fluctuations observed during actual disease outbreaks [8].

In deterministic models, fixed parameters and initial conditions are used to predict the number of individuals in each compartment over time, resulting in predictable and repeatable outcomes for a given set of initial values. These models are effective at generating a single, smooth epidemic trajectory that can aid in understanding the general trend of an outbreak. While typically simpler and computationally efficient, deterministic models often fail to capture the variability present in real-world epidemiological data [14].

These limitations highlight the need for ongoing refinement and adaptation of modeling approaches to reflect local contexts better and improve predictive accuracy. This motivates our focus on stochastic models, which incorporate variability and account for the inherent randomness in disease transmission and recovery processes. Unlike deterministic models, stochastic models can produce different outcomes even under identical initial conditions. Such models are particularly well-suited to capturing the dynamics of the COVID-19 outbreak, where local social, cultural, and political factors can substantially influence individual health behaviors and the effectiveness of interventions.

Access to healthcare in Kenya, as in many African countries, is heavily influenced by socioeconomic status. Individuals with lower income levels often face significant barriers to accessing quality healthcare, as poverty limits their ability to obtain essential health services, medications, nutritious food, and clean water—factors that can exacerbate existing health issues [15]. During the COVID-19 outbreak, misinformation and cultural misconceptions about the virus and vaccines further challenged public health efforts [16]. Cultural norms surrounding health and wellness play a key role in shaping how individuals perceive and respond to health threats [17]. In Kenya, communal living and frequent social gatherings initially made the implementation of social distancing measures particularly difficult [18].

Since the effectiveness of public health responses is often tied to the enforcement of government policies, Kenya’s swift implementation of lockdowns, travel bans, and curfews helped mitigate the spread of COVID-19. These unique social and structural factors influenced how individuals and communities responded to the outbreak, underscoring the importance of a multifaceted and context-sensitive approach to public health.

To address these local dynamics, we adopt a simple stochastic framework based on DSA to transform the classical SIR model into a survival-based stochastic formulation. This approach enables statistical inference through the derivation of a likelihood function that captures the inherent randomness of disease spread [19]. DSA method uses an ODE-derived survival function to represent the probability that an individual remains susceptible up to a certain time, *t*. The hazard function, which represents the rate at which susceptible individuals become infected, is proportional to the number of infectious individuals. The DSA approach allows for a top-down analysis of epidemic data using a mass transfer model based on lumping of the individual-based SIR stochastic model making it easier to perform statistical inference. This approach leverages the principle of propagation of chaos, which simplifies the dynamics in large populations [19]. From the resulting transformed stochastic SIR system, a likelihood function is derived for parameter estimation and model simulation [8, 20].

#### DSA model

Consider the equation (1) with the initial conditions *s*_*t*_ = 1, *ι*_*t*_ = *ρ*, and *r*_*t*_ = 0 The first equation in 1 can be solved using the method of integrating factors and variation of parameters for the second to get

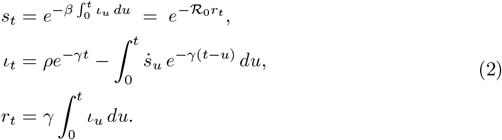

- From the first equation above, we see that *s*_*t*_ may be considered as the probability that a susceptible individual in a very large population remains uninfected beyond time *t*. Specifically, *s*_*t*_ = *P* (*T*_*I*_ *> t*) is the probability that an individual remains susceptible up to time (*t*), where *T*_*I*_ is the time of infection of a randomly chosen initially susceptible individual.
- The hazard function in this approach represents the instantaneous rate at which a susceptible individual *i* becomes infected at time *t*. From the survival function above, the hazard function is defined by *λ*_*t*_ = *βι*_*t*_. The cumulative hazard function represents the accumulated risk of an infection for the susceptible individual *i* occurring up to time *t* and is defined by 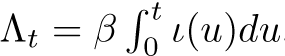.
- The final epidemic size *s*_∞_ can be derived from the survival function to represent the proportion of individuals who survive the epidemic. The probability that *P* (*T*_*I*_ = ∞) = *s*_∞_ defines the limiting probability of remaining susceptible at the end of the epidemic outbreak.

#### Likelihood function

The survival function characterizes the time until an event—such as infection or recovery—occurs. To use it in practice, we require observed infection and removal times for each recorded individual. The corresponding likelihood function, derived from the survival formulation, allows for estimation of the parameters in the SIR model.

In general, the confirmed individual is immediately isolated, so the time of confirmation is used as the time of recovery. Let *T*_*Iw*_ define the random time at which a randomly chosen, initially susceptible individual was confirmed as a COVID-19 positive case, and let *T*_*w*_,*w* = 1, 2, 5 denote the end time point for each wave. Then

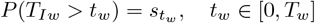

represents the proportion of individuals who remain susceptible at time point *t*_*w*_, thus 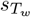 represents the proportion at the end of each wave and *τ*_*w*_ is the final size of the epidemic. Let *ρ*_*w*_ denote the proportion of new cases on the first day of each epidemic wave, *r*_*w*_ = *τ*_*w*_ + *ρ*_*w*_ be the limiting proportion of the recovered individuals, then

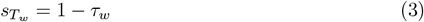

where *τ*_*w*_ ∈ [0, 1) defines the probability that *T*_*Iw*_ *< T*_*w*_, and when *T*_*w*_ = ∞,

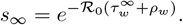

The conditional survival function, conditioned on *T*_*Iw*_ *< T*_*w*_, is given by

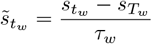

and its probability density is therefore

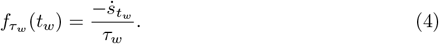

Note that we have 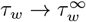 as *t*_*w*_→ ∞ (i.e., when *T*_*w*_ *→ ∞*).

From the second equation in (1)

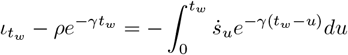

dividing throughout by *τ*_*w*_, we can use the definition of 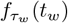 to get the convolution formula

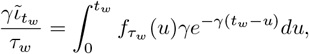

where 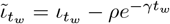 and the density of the variable 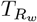 is given by (see, [8, 19, 21])

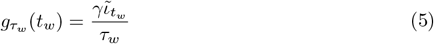

For a completely observed epidemic until time *T*_*w*_, individual disease records obtained have both the infection times *t*_*w,i*_ and removal time *t*_*w,r*_. In such a case, the individual likelihood (that is, one for a single individual) is given by

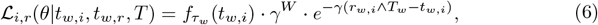

where *W* is the even indicator such that *W* = 0, if *r*_*w,i*_ ∧ *T*_*w*_ = *T*_*w*_, and *W* = 1 otherwise. In a population of *n* individuals with complete records 6 the corresponding likelihood is then the product of the individual likelihoods, that is, the infection events are assumed to be independent.

For the COVID-19 epidemics described above where partial observation results in only the time of removal for the *i*-th individual, the likelihood contribution is

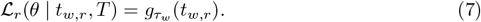

This approach is demonstrated in the data analysis in Section 1.

#### Effective population size

The effective population size refers to the total number of cases divided by the probability of being in the epidemic. In the DSA formulation the equations are in proportions of individuals in each compartment, it is difficult to apply this in real epidemics as the virus usually spreads in a finite population which is not usually known. In the COVID-19 dataset we analyze in this study, only the removal times are recorded, so there is a need to estimate the actual population at risk.

The estimate of effective population size 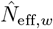 can be computed as

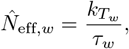

where

- 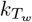 is the count of observed infected individuals over the time horizon *T*_*w*_ and *τ*_*w*_ for each of the waves.
- From the above computations, the final epidemic size for the combined three waves 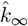 is given by 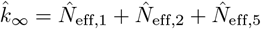.

## Data analysis and model fitting

Kenya is one of the major economies in the Eastern African region with a total population of about 50 million in the year 2020. The country has a county government structure with 47 counties, over 9000 health facilities that include public, private and faith-based organizations. The healthcare worker to population stood at about 4 workers for every 10,000 citizens [17, 22].

### COVID-19 situation report and case classification

The first case of COVID-19 virus was confirmed on 13th March 2020. part of the country’s efforts to control the spread of COVID-19 during the early stages of the pandemic, an international travel ban was imposed on March 25, 2020, and lasted until August 1, 2020, when international flights resumed.

As with other diseases, cases of COVID-19 were broadly defined under a three-level system: suspected cases are those who showed clinical signs and symptoms of having COVID-19, but had not been laboratory-tested. A probable case included patients with COVID-19 symptoms and has either been in close contact with a positive case, or is in an area particularly affected by COVID-19. Then there were confirmed cases that is, a person with laboratory confirmation of COVID-19 infection.

The cases were reported by international organizations such as the WHO and European CDC using reported case figures submitted by Kenyan government. Efforts were made report confirmed cases wherever possible because laboratory confirmation gave international organizations a higher degree of certainty.

The long reporting chain of reporting a COVID-19 case involved diagnosis by a doctor or laboratory, submission to the local health department, recording in the reporting system, aggregation by governmental organizations, and collation by international bodies like WHO or European CDC. This chain could take several days, so the figures reported on any given date do not necessarily reflect the number of new cases on that specific date, so number of actual cases is higher than the number of confirmed cases.

To understand the scale of the COVID-19 outbreak and how it progressed, it is important for scientific research to determine how many people are infected by COVID-19. Since the actual number of COVID-19 infections is not known, we would like to estimate it from data using the simple DSA model. Kenya faced several challenges during the outbreak like many other African countries and therefore struggled to cope with the runaway transmission. Among them is limited testing capacity, which made it difficult to test large numbers of people who were exposed or showed symptoms of the virus. A significant portion of the population has no income and therefore had no access to health insurance, with the high cost of the test, many people could not afford COVID-19 to take the test, and the subsidized or free testing was limiting to only a few hots-spot areas and groups of workers in sensitive public service. There is a wide regional differences in testing rates, with urban and metropolitan areas had better access to testing facilities than others. Awareness campaigns were done to reach the masses to enhance self protection and access to treatment and quarantine facilities. However the uneven distribution of mobile digital access across counties hampered effective communication and coordination for testing, treatment and quarantine.

These challenges highlight the need for better preparation on future epidemics so that targeted interventions to enhance testing capacity, access to timely and effective treatment, and therefore save lives lost unnecessarily. As of the latest available data, Kenya reported 5,689 confirmed deaths from COVID-19.

As part of the greater goal of controlling the virus spread and collect data more effectively, the “Jitenge” (a Swahili word meaning “quarantine yourself”) system was designed and it became a crucial part of Kenya’s response to the COVID-19 pandemic. It was part of broader public health collaborations between the Government of Kenya and CDC Kenya. The aim of the system was to support patients and enhance surveillance. The “jitenge” system was used to monitor the health status of individuals, particularly travelers entering Kenya. It allowed individuals, including truck drivers, to report symptoms and receive test results after entry into Kenya. The system supported COVID-19 contact tracing efforts, helping to identify and manage potential cases and facilitated the collection and management of health data, which was crucial for tracking the spread of the virus. The system was integrated into national and county-level emergency operations centers to streamline response efforts. The system provided a platform for training and mentoring healthcare workers on infection prevention and control, as well as “Jitenge” enabled real-time monitoring of health trends and facilitated timely interventions [15, 23].

We illustrate the DSA model using data from Kenyan COVID-19 outbreak records, which is publicly available at Our World in Data, by analyzing records for the first two waves and the fifth wave of the outbreak.

The recorded case counts were randomly distributed across each day to approximate a continuous time-to-event distribution. Since testing was conducted in healthcare facilities, the day of a positive test may be regarded as a surrogate for the time of removal. Consequently, the exact timing of symptom onset is less relevant. For each record, we consider then an onset event unknown and consider only a corresponding removal time. We note that once symptoms were confirmed, patients were hospitalized or quarantined, effectively preventing further transmission.

To fit the Kenya COVID-19 dataset using the DSA approach, we approximate posterior densities using the Hamiltonian Monte Carlo (HMC) sampler [24], in Rstan library, an open-source statistical package for R software [25, 26].

In this analysis, the COVID-19 data fall under the *partially observed likelihood* setting described in Section 1, corresponding to the case where only removal times are recorded (cf. (7)). The available individual-level records consist of COVID-19 confirmation dates, which in practice triggered quarantine—either in hospital or at home. During the quarantine period, we assume that infected individuals were no longer infectious, as contact with the population at risk was effectively eliminated. Consequently, from the point of view of the SIR model (1), *these confirmation dates are treated as removal (recovery) times* in the model. The posterior parameter estimates obtained from fitting the model to the data for the three selected epidemic waves are presented in Tables 1, 2, and 3.

**Table 1.** Wave 1 Posterior Summary Statistics. Posterior summaries for key model parameters including transmission rate (*β*), removal rate (*γ*), the initial infected count (*ρ*), the basic reproduction number (ℛ_0_), and the effective population size *N*_eff,1_.

| Statistic | $\beta$ | $\gamma$ | $\rho$ | $\mathcal{R}_0$ | $N_{\text{eff},1}$ |
| --- | --- | --- | --- | --- | --- |
| Min | 0.3679 | 0.3078 | 0.0000100 | 1.1158 | 449916 |
| LCL (2.5%) | 0.4226 | 0.3656 | 0.0000101 | 1.1405 | 537078 |
| Median | 0.4387 | 0.3820 | 0.0000103 | 1.1482 | 561474 |
| Mean | 0.4388 | 0.3823 | 0.0000105 | 1.1485 | 563365 |
| UCL (97.5%) | 0.4538 | 0.3980 | 0.0000107 | 1.1565 | 587210 |
| Max | 0.5141 | 0.4607 | 0.0000142 | 1.1952 | 691491 |
| SD | 0.0227 | 0.0236 | 5.144e-07 | 0.0116 | 36440 |

**Table 2.**
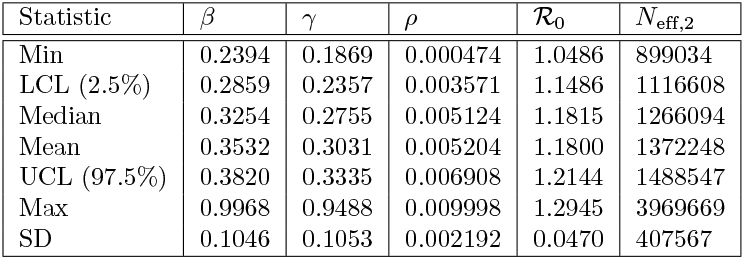
Wave 2 Posterior Summary Statistics. Posterior summaries for key model parameters including transmission rate (*β*), removal rate (*γ*), the initial infected count (*ρ*), the basic reproduction number (ℛ_0_), and the effective population size *N*_eff,1_.

**Table 3.** Wave 5 Posterior Summary Statistics. Posterior summaries for key model parameters including transmission rate (*β*), removal rate (*γ*), the initial infected count (*ρ*), the basic reproduction number (ℛ_0_), and the effective population size *N*_eff,5_. Note the relatively high value of (ℛ_0_) compared to the previous waves.

| Statistic | $\beta$ | $\gamma$ | $\rho$ | $\mathcal{R}_0$ | $N_{\text{eff},5}$ |
| --- | --- | --- | --- | --- | --- |
| Min | 0.3876 | 0.1031 | 0.00308 | 2.665 | 229742 |
| LCL (2.5%) | 0.4097 | 0.1160 | 0.00439 | 3.299 | 233106 |
| Median | 0.4167 | 0.1204 | 0.00482 | 3.455 | 234587 |
| Mean | 0.4171 | 0.1206 | 0.00487 | 3.471 | 234787 |
| UCL (97.5%) | 0.4237 | 0.1249 | 0.00533 | 3.631 | 236202 |
| Max | 0.4522 | 0.1511 | 0.00726 | 4.263 | 247709 |
| SD | 0.0101 | 0.0065 | 0.00069 | 0.241 | 2294 |

Using the parameter estimates for each wave, we generated posterior density plots for the model parameters, including the basic reproduction number and the effective population size. To illustrate the Bayesian sampling procedure, we employed two chains of 4,000 iterations each, discarding the first 1,000 iterations as burn-in. The resulting trace plots display the evolution of sampled values across iterations, confirming both the stability of the sampling process and the reliability of the estimates. Finally, for each wave, we present the model fit obtained from the corresponding parameter estimates, overlaid on the observed case histograms.

### Waves 1 and 2

The corona virus outbreak in Kenya begun on 5th March 2020 when the first case was confirmed, with a peak on 7th August 2020 with 670 cases and ended on 18th September 2020. During this period, the government had put in place strict protocols and restrictions to curb the spread of the disease. Schools were closed, movement from the city to the rural areas were restricted and a system of contact tracing was in place to ensure those who get exposed were traced, tested and treated on time. The overwhelmed health facilities did the best they can to assist patients under their care. The spread therefore slowed down and the peak took a longer period compared to the other waves.

The second wave began on 1st October 2020 with 184 cases, with a peak on 30th November 2020 when 1100 cases, and ended on 2nd February 2021, taking a total of 120 days.

Starting with the analysis of results for wave 1, we see in Fig 1 the histogram of the number of new cases for the 200 days of wave 1. This is the trend we hope to capture in our model fit. The fit is done using Rstan in Rn((version 4.4.2 (2024-10-31)) and the resulting posterior parameter estimates are shown in Table 1. These metrics provide a robust characterization of central tendency, dispersion, and uncertainty, supporting inference and model interpretation. Notably, the skewness observed in *ρ* is reflected in the asymmetry between its percentiles and wider credible interval.

**Fig 1.**
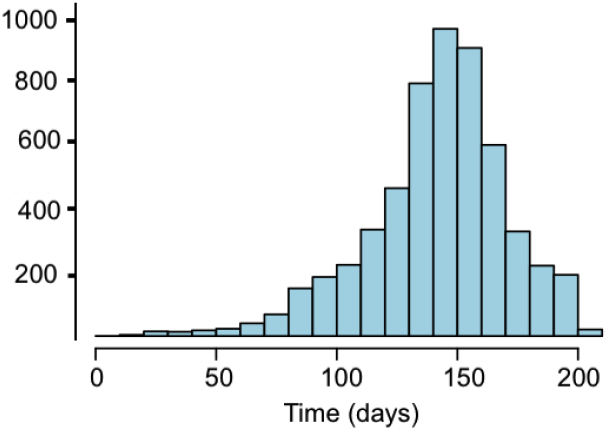
Wave 1 Incidence. The histogram shows the number of daily confirmed COVID-19 cases per day for COVID-19 cases in Kenya during the first wave. The recordings were compiled from various health facilities each day. Source: Our World in Data

The convergence of the four chains used in the HMC sampling with the density plots is shown in Fig 2 confirming the stability of the posterior density estimates and HMC Chain Diagnostics for *β, γ*, ℛ_0_, and *ρ*. The left panel depicts posterior distributions for the transmission rate (*β*), removal rate (*γ*), and basic reproduction number (ℛ_0_ = *β/γ*) that exhibit smooth, symmetric, bell-shaped curves consistent with normality, indicating well-constrained estimates and high confidence in the inferred epidemic parameters. In contrast, the posterior distribution of *ρ*, the initial number of infected individuals at the start of the modeling period, shows pronounced right skewness, reflecting uncertainty in early outbreak conditions and the potential for under-reported cases prior to observation. The right panel depicts the trace diagrams for four HMC sampling chains demonstrate good mixing and convergence across parameters. The initial 1,000 iterations, shaded in gray, were discarded as burn-in to ensure posterior samples reflect the stationary distribution.

**Fig 2.**
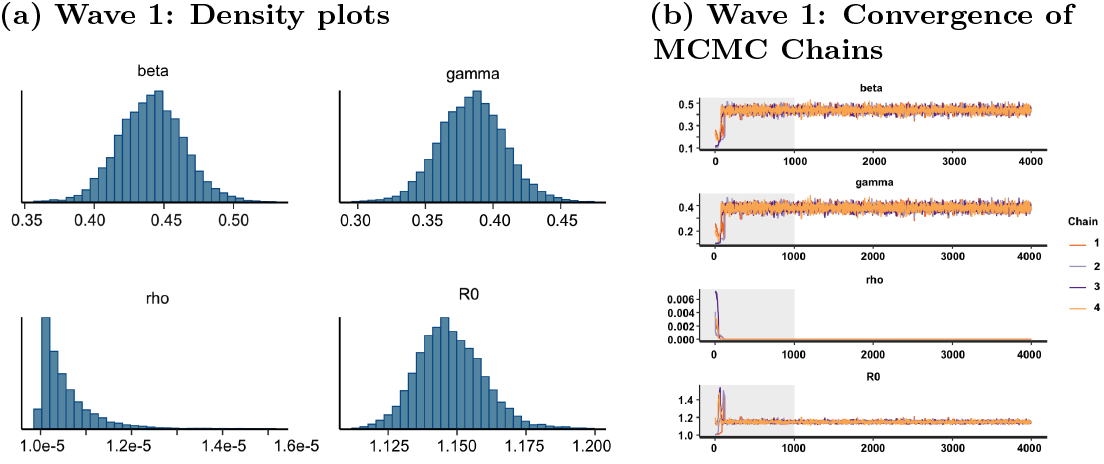
Wave 1 Analysis. (a): Posterior Density Estimates and HMC Chain Diagnostics for *β, γ*, ℛ_0_, and *ρ*. The left panel depicts posterior distributions for the transmission rate (*β*), removal rate (*γ*), and basic reproduction number (ℛ_0_ = *β/γ*) exhibit smooth, unimodal curves, indicating well-constrained estimates and high confidence in the inferred epidemic parameters. (b): Diagnostic trace plots with 1000 burn-in steps (shared region) indicate well–mixing chains.

DSA model fit that follows the cumulative cases over the period in Fig 3 bounded by the 95% credible interval. The close alignment between observed and modeled recoveries throughout most of the wave reflects strong parameter identifiability and robust inference, as supported by well-mixed HMC chains and symmetric posterior densities for *β, γ*, ℛ_0_, and *ρ*. The right-skewed posterior of *ρ* captures uncertainty in early outbreak conditions. The observed overestimation near the end of the wave likely stems from reporting artifacts, where some patients may have been counted multiple times.

**Fig 3.**
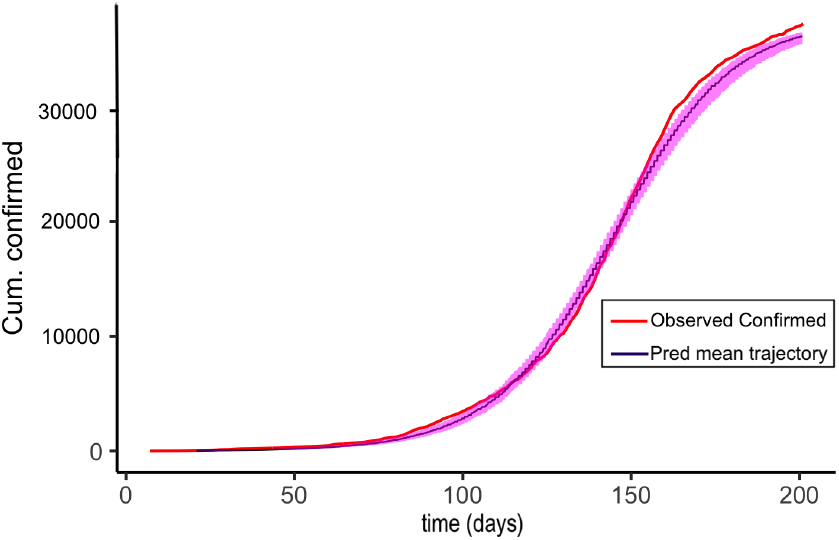
Wave 1 DSA Model Fit. Model fit for wave 1 of the outbreak, showing cumulative confirmed cases hospitalizations (that is, removal times) in red, overlaid with the Dynamical Survival Analysis model output in blue. The model trajectory is based on the computed means of posterior parameter estimates (*β, γ*, ℛ_0_, and *ρ*), with a 95% confidence interval shaded in magenta.

We next turn to the analysis of data from wave 2, carried out in a manner analogous to that of wave 1. The number of newly confirmed cases is presented as a histogram in Figure 4, based on the referenced data source. As in the first wave, we performed HMC sampling with four chains; the corresponding convergence diagnostics and posterior density plots are shown in Figure 5. The parameter estimates are reported in Table 2. Finally, using these posterior estimates, we overlay the DSA curve on the histogram, yielding the fitted results in Figure 6.

**Fig 4.**
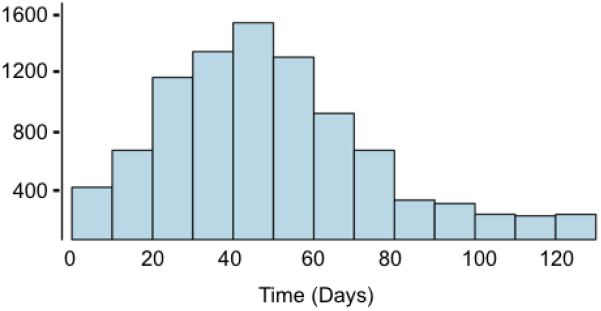
Wave 2 Incidence. The histogram shows the number of daily confirmed COVID-19 cases per day for COVID-19 cases in Kenya during the second wave. The recordings were compiled from various health facilities each day Source: Our World in Data

**Fig 5.**
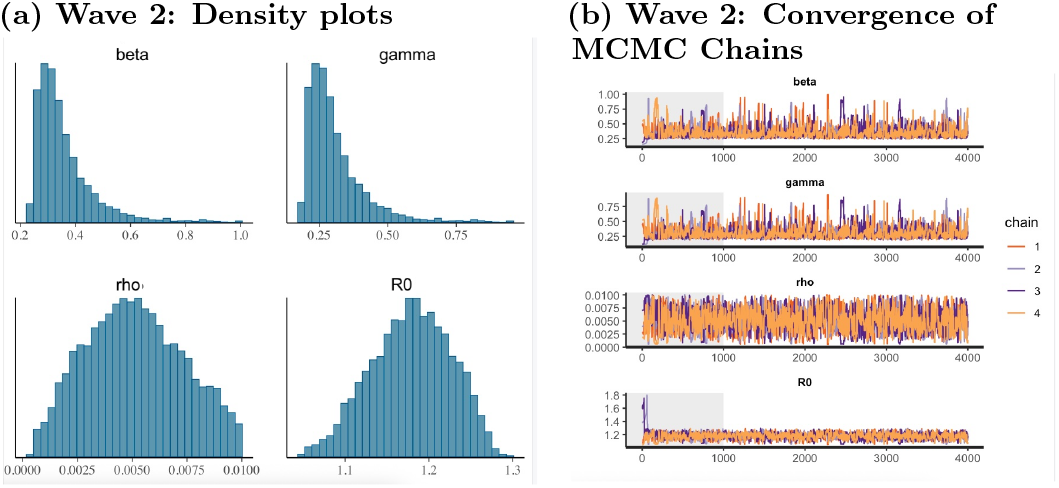
Wave 2 Analysis. (a): Posterior Density Estimates and HMC Chain Diagnostics for *β, γ*, ℛ_0_, and *ρ*. The left panel depicts posterior distributions for the transmission rate (*β*), removal rate (*γ*), and basic reproduction number (ℛ_0_ = *β/γ*) exhibit smooth, unimodal curves. Note the skewness of the posterior distributions of *β* and *γ* that is markedly different from their analogues in wave 1. (b): Diagnostic trace plots with 1000 burn-in steps (shared region) indicate well–mixing chains.

**Fig 6.**
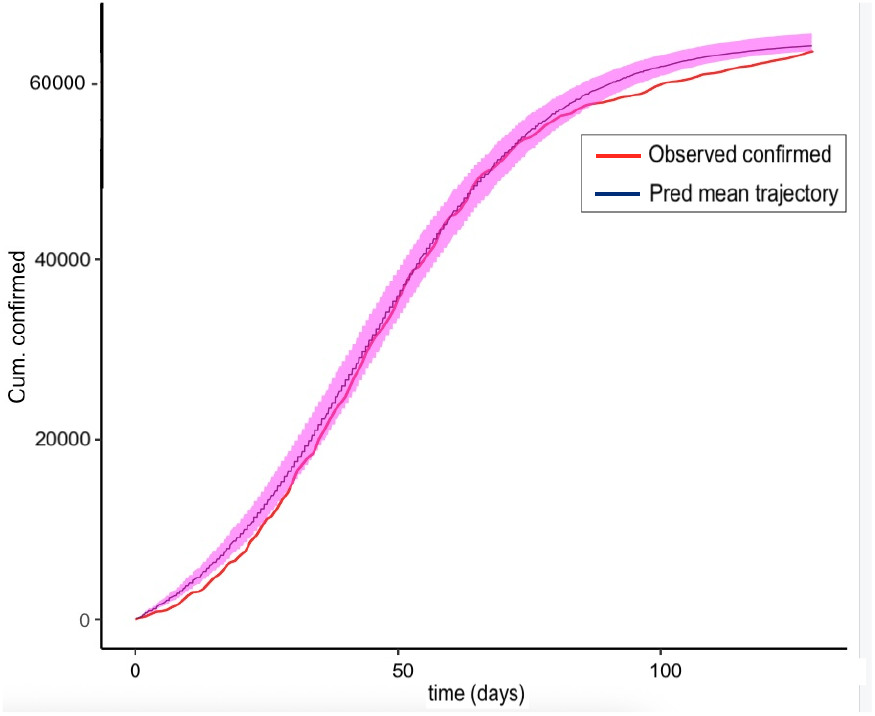
Wave 2 DSA Model Fit. Model fit for wave 2 of the outbreak, showing cumulative confirmed cases hospitalizations (that is, removal times) in red, overlaid with the Dynamical Survival Analysis model output in blue. The model trajectory is based on the computed means of posterior parameter estimates (*β, γ*, ℛ_0_, and *ρ*), with a 95% confidence interval shaded in magenta. Lack of fit in the initial and final phase of the wave may indicate under-reporting of cases.

### Wave 5 - The highest COVID peak in Kenya

The highest wave of COVID19 in Kenya was recorded between December 6, 2021, to April 2, 2022 caused by the highly infectious Omicron mutation of COVID-19. The infections peaked on December 29 2021 when over 3000 new confirmations were recorded. December is a holiday month in Kenya with family gatherings in the villages. This increased social interactions during the holiday season likely contributed to the spike in cases. At the same, time emergence of the highly transmissible Omicron variant led to a rapid increase in infections. Another reason may be the reduced Adherence to Protocols due to fatigue from prolonged restrictions and closures, which were relaxed during this season. The government escalated the vaccine mass education campaign, improved public health surveillance, testing, and contact tracing effort, which led to an increased vaccination uptake, hence helped reduce the number of new infections. After the first four waves, it is believed that the population developed natural immunity from previous infections, thus reducing infectivity and disease severity [17]. These and other unknown factors led to the rise and fall of daily confirmed cases in the country.

The observed COVID-19 data histogram is shown in Fig 7. Convergence of the MCMC chains and density plots in Fig 8 show the posterior distributions for the transmission rate (*β*), removal rate (*γ*), and basic reproduction number (ℛ_0_ = *β/γ*) alongside that of the initial number of infected individuals (*ρ*). The distributions of the parameters exhibit smooth, symmetric, bell-shaped curves consistent with normality, indicating well-constrained estimates and high confidence in the inferred epidemic parameters.

**Fig 7.**
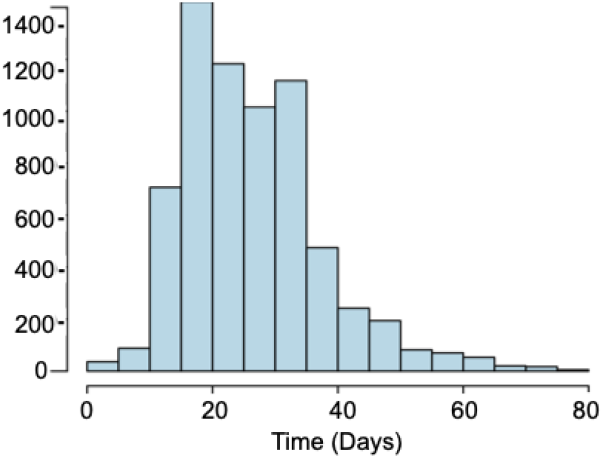
Wave 5 Incidence. The histogram shows the number of daily confirmed COVID-19 cases per day for COVID-19 cases in Kenya during the steep fifth (Omicron) wave. The recordings were compiled from various health facilities each day Source: Our World in Data

**Fig 8.**
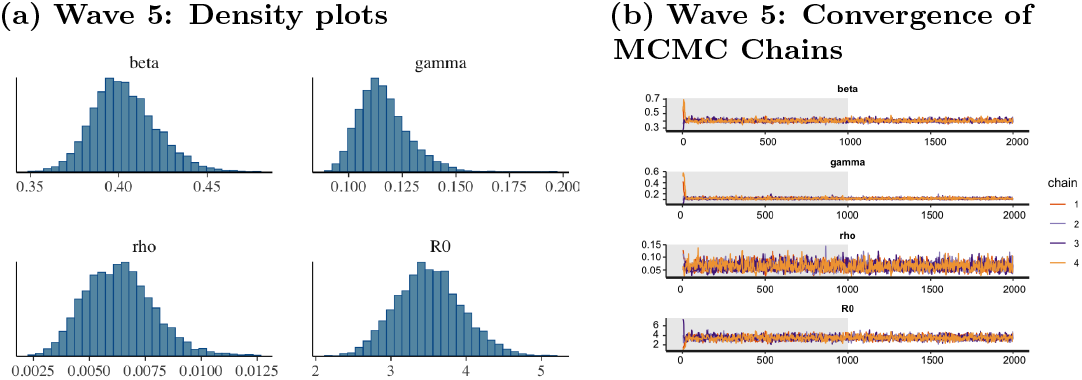
Wave 5 Analysis. (a): Posterior Density Estimates and HMC Chain Diagnostics for *β, γ*, ℛ_0_, and *ρ*. The left panel depicts posterior distributions for the transmission rate (*β*), removal rate (*γ*), and basic reproduction number (ℛ_0_ = *β/γ*) exhibit smooth, unimodal curves. Note that this time the posterior distributions appear more symmetric that in previous waves. (b): Diagnostic trace plots with 1000 burn-in steps (shared region) indicate well–mixing chains.

Besides the density plots, trace diagrams for four HMC sampling chains demonstrate good mixing and convergence across parameters. The initial 1,000 iterations, shaded in gray, were discarded as burn-in to ensure posterior samples reflect the stationary distribution. The estimated posterior parameter values are shown in Table 3.

Model fit for wave 5 of the outbreak, showing cumulative removals (i.e., hospitalizations) in red, overlaid with the Dynamical Survival Analysis model output in blue. The model trajectory is based on the computed means of posterior parameter estimates (*β, γ*, ℛ_0_, and *ρ*), with a 95% credible interval shaded in magenta. The fit closely aligns with observed recoveries across the entire wave, indicating excellent predictive performance and strong parameter identifiability. Posterior density functions for all parameters exhibit smooth, symmetric, bell-shaped curves consistent with normality, and are accompanied by small standard deviations—suggesting high precision and low uncertainty in the estimates. These results reflect both the quality of the data and the stability of the inference process.

**Fig 9.**
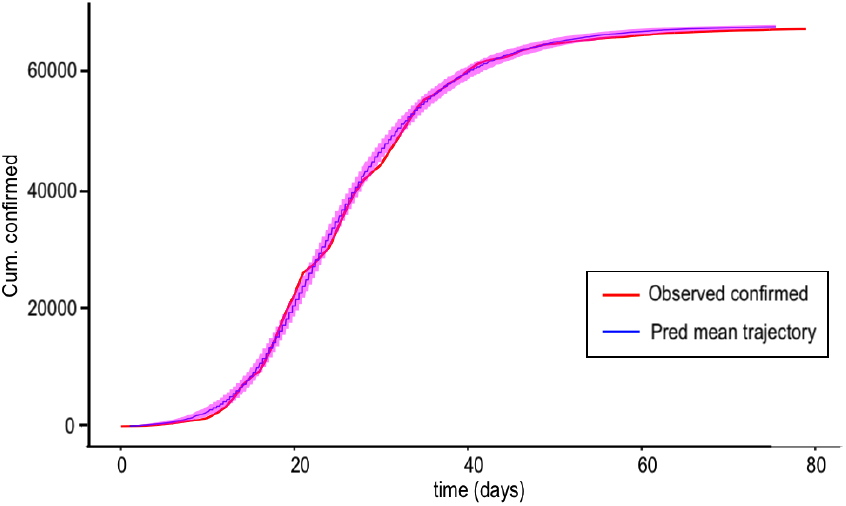
Wave 5 DSA Model Fit. Model fit for Wave 1 of the outbreak, showing cumulative confirmed cases hospitalizations (that is, removal times) in red, overlaid with the Dynamical Survival Analysis model output in blue. The model trajectory is based on the computed means of posterior parameter estimates (*β, γ*, ℛ_0_, and *ρ*), with a 95% confidence interval shaded in magenta. Lack of fit in the initial and final phase of the wave may indicate under-reporting of cases.

## Discussion

This paper presents a comprehensive illustration of the Dynamical Survival Analysis (DSA) approach, using data from the 2020–2022 COVID-19 outbreak in Kenya. The DSA framework is a robust tool for analyzing disease progression in populations, combining survival analysis techniques with dynamical systems theory. This approach facilitates statistical inference and the modeling of epidemic dynamics, particularly in large populations.

Reformulating the SIR model via DSA yields a system of equations that captures the stochastic nature of disease transmission. This transformation enables the derivation of survival and hazard functions, as well as the likelihood function required for Bayesian inference and parameter estimation, thereby streamlining the model-fitting process to real epidemiological data. The resulting model is conceptually simple, computationally efficient, and well-suited for analysis with real-world data. It addresses the complexity found in many COVID-19 models while enabling accurate predictions of disease trajectories (see related DSA-based approaches in [8, 19]).

In this study, the DSA method is applied to COVID-19 outbreak data from Kenya, capturing the nationwide spread between 2020 and 2022. The country experienced five major waves, with daily confirmed case counts for each wave shown in Fig 1, 4, 7. For illustrative purposes, we focus on Wave 1, Wave 2, and Wave 5, with the corresponding estimated parameter values presented in Tables 1, 2, and 3, respectively.

A key goal of our analysis was to estimate the basic reproduction number and the underlying epidemiological rates governing disease transmission and recovery across successive waves of the COVID-19 epidemic in Kenya. The infection rate (*β*) and removal rate (*γ*) varied substantially across regions and epidemic contexts. In Kenya, as in many other African countries, previous modeling studies have reported infection rates typically ranging from 0.2 to 0.5 and recovery/removal rates from 0.1 to 0.2 [27]. At the global level, estimates from the World Health Organization (WHO) suggest *β* values between 0.3 and 0.6 and *γ* values between 0.1 and 0.3 [15]. Our posterior estimates of *β, γ*, and the basic reproduction number (ℛ_0_) align well with these previously reported ranges.

Because 1*/γ* represents the average duration of the infectious period, our findings indicate that during wave 1 and wave 2 patients were infectious for approximately 2–3 days, whereas in wave 5 the infectious period extended to about 7 days. This longer duration in wave 5 coincided with a higher epidemic peak, prolonged illness, and potentially more severe complications. The corresponding ℛ_0_ estimates reinforce this pattern: in wave 5 each infectious individual was estimated to generate about three secondary cases, compared with only one secondary case in wave 1 and wave 2. It is important to note that the total population at risk and the duration of each wave also influenced these differences. The parameter *ρ*, distinct from ℛ_0_, represents the number of confirmed new cases at the onset of each epidemic wave rather than the initial proportion of infected individuals.

The epidemic trajectory in wave 5, as captured by the SIR model fitted with MCMC under the DSA framework, closely reproduces the observed dynamics. The estimated ℛ_0_ for wave 5 is more than double the values observed in wave 1 and wave 2, underscoring the heightened transmission intensity due to the properties of the Omicron COVID-19 mutation. These results highlight the capacity of the DSA approach to provide reliable inference, with posterior uncertainty quantification further enhancing prediction robustness. Spillover of infected individuals from earlier waves may also have contributed to the observed dynamics in later waves.

In summary, the results presented here highlight the utility of DSA as a principled framework for integrating classical epidemic models with real-world data. This approach allows for the incorporation of stochastic epidemic dynamics within a likelihood-based inference framework, providing both robust parameter estimation and reliable forecasting. The integration of DSA into epidemic modeling offers valuable theoretical insights and practical tools for understanding disease transmission—particularly in settings characterized by heterogeneous data quality and limited surveillance infrastructure. As such, DSA holds considerable promise for informing more adaptive and effective public health interventions in future epidemic scenarios.

Within the African context, including Kenya, estimates of ℛ_0_ have shown substantial heterogeneity, reflecting variations in healthcare capacity, population density, public health interventions, and socioeconomic conditions. Reported estimates across Africa range from 1.98 to 9.66, with a median of about 3.67. By contrast, global estimates suggest that ℛ_0_ converged to approximately 4.5 during the early phase of the pandemic, before widespread control measures were implemented [28].

## Author Contributions

GAR and JW developed the model, JW and BC analyzed the data. GAR and JW drafted the initial manuscript. AG contributed to data analysis, manuscript revisions, and the development of the necessary software scripts. All authors reviewed and approved the final version of the manuscript.

## Data Availability and Supplementary Materials

The dataset is publicly available at https://ourworldindata.org/coronavirus, the code used in MCMC analysis, and an appendix of the MCMC chains output are publicly available via;

## Acknowledgment

The work of GR and JW was partially funded by the HEALMOD initiative at The Ohio State University. JW was partially funded by IMU-CDC.

